# COVID-19: Forecasting short term hospital needs in France

**DOI:** 10.1101/2020.03.16.20036939

**Authors:** Clément Massonnaud, Jonathan Roux, Pascal Crépey

## Abstract

Europe is now considered as the epicenter of the SARS-CoV-2 pandemic, France being among the most impacted country. In France, there is an increasing concern regarding the capacity of the healthcare system to sustain the outbreak, especially regarding intensive care units (ICU). The aim of this study was to estimate the dynamics of the epidemic in France, and to assess its impact on healthcare resources for each French metropolitan Region. We developed a deterministic, age-structured, Susceptible-Exposed-Infectious-Removed (SEIR) model based on catchment areas of each COVID-19 referral hospitals. We performed one month ahead predictions (up to April 14, 2020) for three different scenarios (*R*_0_ = 1.5, *R*_0_ = 2.25, *R*_0_ = 3), where we estimated the daily number of COVID-19 cases, hospitalizations and deaths, the needs in ICU beds per Region and the reaching date of ICU capacity limits. At the national level, the total number of infected cases is expected to range from 22,872 in the best case (*R*_0_ = 1.5) to 161,832 in the worst case (*R*_0_ = 3), while the total number of deaths would vary from 1,021 to 11,032, respectively. At the regional level, all ICU capacities may be overrun in the worst scenario. Only seven Regions may lack ICU beds in the mild scenario (*R*_0_ = 2.25) and only one in the best case. In the three scenarios, Corse may be the first Region to see its ICU capacities overrun. The two other Regions, whose capacity will be overrun shortly after are Grand-Est and Bourgogne-Franche-Comté. Our analysis shows that, even in the best case scenario, the French healthcare system will very soon be overwhelmed. While drastic social distancing measures may temper our results, a massive reorganization leading to an expansion of French ICU capacities seems to be necessary to manage the coming wave of critically affected COVID-19 patients.

## 2 Introduction

On December 31, 2019, Chinese authorities informed WHO of grouped pneumoniae cases^1^. The majority of these cases were linked the Huanan South China Seafood Market, in the city of Wuhan, Hubei province, China. A novel coronavirus, SARS-Cov-2, was identified on January 7, 2020, as the cause of this outbreak. On January 13, the first case outside of China was confirmed in Thailand^2^. The first cases on the European continent were confirmed in France on January 24^3^. The World Health Organization (WHO), declared the outbreak a Public Health Emergency of International Concern on 30 January 2020^4^, and announced a name for the disease on February 11: COVID-19^5^.

As of March 5, the European Centre for Disease Prevention and Control (ECDC) reported 91,315 COVID-19 confirmed cases in 81 countries, and 3,282 deaths (3.4%). In Europe, 38 countries reported cases, Italy accounting for the majority of them, with 3,089 cases out of 4,290 (72%), and 107 deaths out of 113 (94.7%). France was ranked second with 423 cases and 5 deaths (1.2%)^6^. On March 11, WHO declared a pandemic, as 106 countries reported 118,628 confirmed cases and 4,292 deaths (3.6%)^7^. In Europe, the number of countries affected increased to 47, Italy still accounting for the majority of cases.^8^

Figure 1 shows the number of cases in metropolitan France from January 22 to March 14 (source Santé Publique France). As of March 10, midnight, 2,030 cases were confirmed, leading to 44 deaths (2.2%)^8^. The two Regions the most impacted were Grand Est and Ile-de-France, with 489, and 468 cases, respectively. People aged over 75 years accounted for 19% of the cases but around 75% of the deaths^8^. As of March 10, 102 cases had been hospitalized into Intensive Care Units (ICU), 38% of them aged 65 years or less. The doubling time is approximately 72h, as the number of cases increased from 1,126 to 2,269 between March 8 and March 11, and from 2,269 to 4,469 between March 11 and March 14. It must be noted that it is likely that the number of confirmed cases is underestimating the true number of cases as all cases are not necessarily identified due to logistical issues in some Regions^8^.

**Figure 1:**
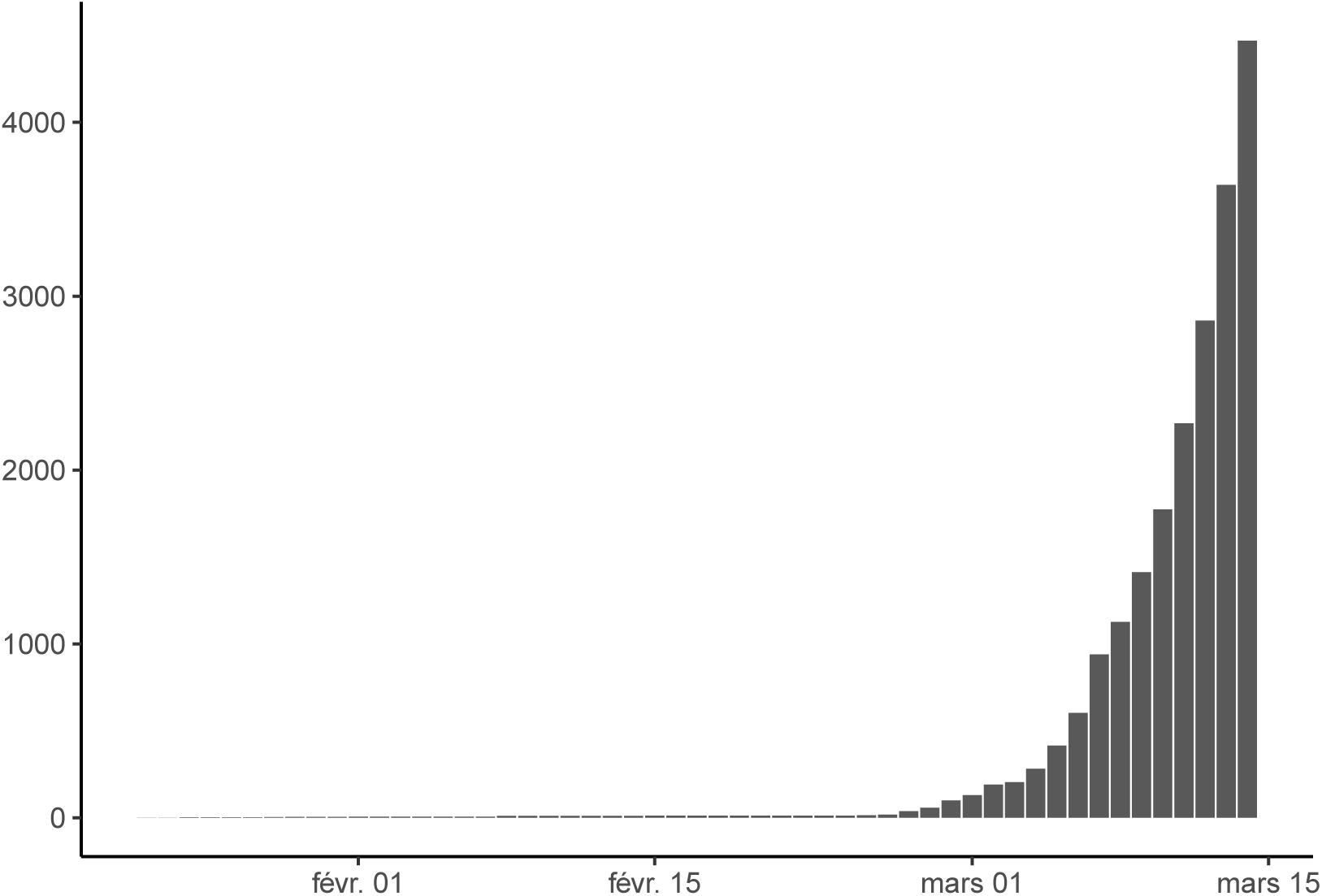
Number of confirmed COVID-19 cases in France from January 22 to March 14

SARS-CoV-2 is a zoonotic virus, and bats are believed to be its reservoir. However, human-to-human transmission has been largely reported^9^. Transmission occurs mostly via droplets and fomites during close unprotected contact. Information about the main transmission parameters is still relatively scarce, and can vary according to the settings, the data, and the methods used.

The basic reproductive number (*R*_0_) was estimated in various studies^9–19^ starting at 1.4, up to 7.23. Most countries have now implemented control strategies involving contact tracing, quarantine, and isolation measures, which are likely to significantly reduce the *R*_0_. Li and colleagues used individually documented case descriptions from China to estimate a reproductive number in the context of control measures (*R*_*c*_)^17^. It was estimated to be 1.54. Zhou and colleagues^19^ estimated a controlled reproduction number between 1.46 and 2.99. They also estimated mean incubation and contagious periods of 5 and 11 days, respectively.

Abbott and colleagues estimated the reproduction number of 24 countries using publicly available data with a 7-day sliding window. Their estimate for France in date of March 13 was between 1.4 and 3.2.^20^

Several models have already been developed to forecast propagation dynamics of COVID-19 epidemics in various settings. However, models in European context are lacking. Danon and colleagues explored disease transmission in England and Wales by adapting an existing stochastic, metapopulation model.^21^ Pinotti and colleagues modeled international importation of COVID-19 cases to assess rapidity of isolation, effect of intervention, and undetection rates.^22^ To this date, no models have been developed to analyze COVID-19 propagation dynamics in France.

## 3 Objective

The objective of this study was to estimate the number of COVID-19 cases, hospitalizations and deaths in France, and to assess the impact of the epidemic on healthcare resources, by estimating the number of required hospital beds in ICU throughout the epidemic, for each region. As a sensitivity analysis and to assess the potential impact of large scale control measures, we varied the *R*_0_ of the epidemic from 3 to 1.5.

## 4 Methods

We listed the 138 hospitals that are identified as referral centers for the treatment of COVID-19 cases in France. Of those, 33 hospitals are listed as primary referral centers, and 98 as secondary referral centers in metropolitan France and 7 are overseas hospitals (not included). We then divided metropolitan France into hospital catchment areas around these referral hospitals using Voronoi polygons (Figure 2). Population structure was inferred for each catchment area from 2016 and 2017 census data provided by the French National Institute of Statistics and Economic Studies (Insee)^23^. Catchment areas were then aggregated by metropolitan Regions [13 French administrative areas with an averaged population of 4.75 millions ranging from 300,000 (Corse) to 12.55 millions (Ile-de-France)]. Data on ICU beds capacity per French Region were retrieved from the “Statistique Annuelle des Etablissements de Santé” (SAE)^24^.

**Figure 2:**
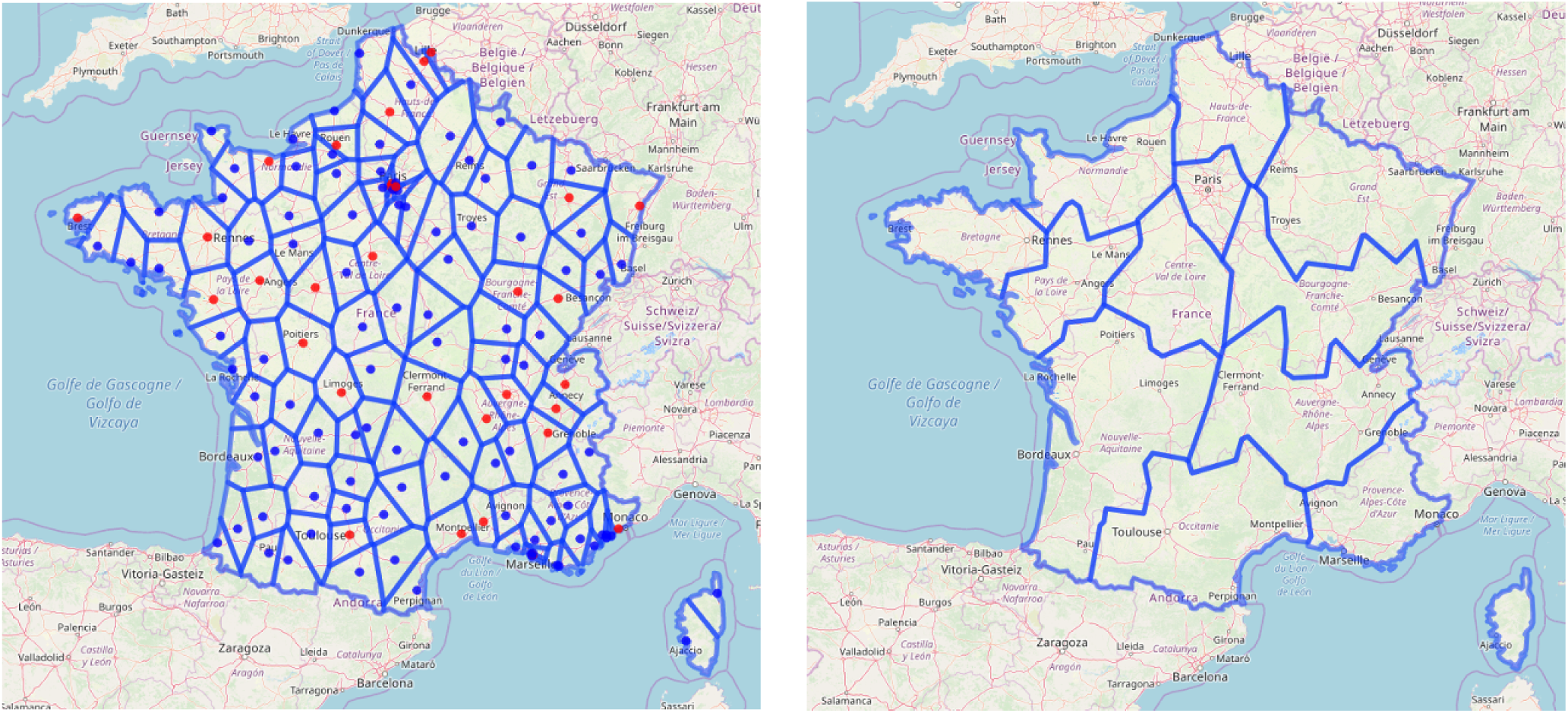
Maps of France divided into hospital catchment areas, then aggregated by French Region.

We developed a deterministic, age-structured, compartmental, Susceptible-Exposed-Infectious-Removed (SEIR) model, as a set of differential equations (Figure 3). The population was divided into 17 age-groups (5 years age-band), and can be either susceptible (S), exposed to the virus but not infectious (E), infected and infectious (I), or removed from the chains of infection (R). We used an inter-individual contacts matrix for the French population estimated by Prem and colleagues^25^ to simulate age-dependent mixing.

**Figure 3:**
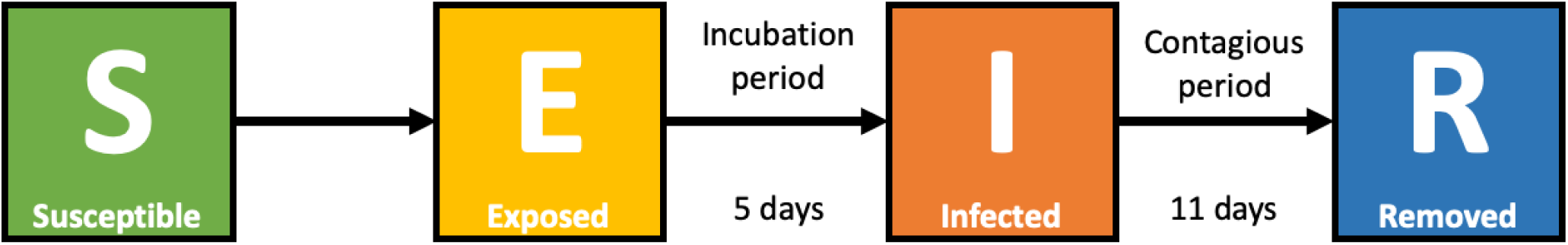
Diagram of the age-structured SEIR model.

We considered an incubation period of 5 days and a contagious period of 11 days^19^. We assumed an average length of stay in hospitals and in ICU wards of 15 days for all patients, of all ages. We estimated the age-dependent susceptibility of the population based on the age distribution of infected cases reported by Wu and colleagues in China^11^. We standardized the Chinese age distribution of cases to the French population to estimate the expected age distribution of cases for France. We then fitted the age dependent susceptibility vector using the R implementation of the subplex algorithm provided by the nloptr package.

Based on the estimated number of new cases per day, we inferred different outcomes. First, we computed the number of severe cases, defined according to Guan and colleagues^18^. We used their estimations of age-dependent severity risks across four age groups, which were dispatched across our 17 age groups (Table 1). We also estimated the number of cases which will require hospitalization to ICU using age-dependent risks from Guan and colleagues^18^ (Table 1). Using data from Yang and colleagues^26^, we set the proportion of cases which will require mechanical ventilation to 0.711 (not age-dependent). The number of deaths was estimated using age-dependent deaths risks (10 age groups) provided by the Italian National Institute of Health (Istituto Superiore di Sanità) (Table 1)^27^.

**Table 1:** Parameters used to compute the main outcomes.

| Age group | Severity risk | ICU risk | Death risk |
| --- | --- | --- | --- |
| 0-4 | 0.11 | 0.02 | 0.000 |
| 5-9 | 0.11 | 0.02 | 0.000 |
| 10-14 | 0.11 | 0.02 | 0.000 |
| 15-19 | 0.12 | 0.02 | 0.000 |
| 20-24 | 0.12 | 0.02 | 0.000 |
| 25-29 | 0.12 | 0.02 | 0.000 |
| 30-34 | 0.12 | 0.02 | 0.002 |
| 35-39 | 0.12 | 0.02 | 0.002 |
| 40-44 | 0.12 | 0.02 | 0.002 |
| 45-49 | 0.12 | 0.02 | 0.002 |
| 50-54 | 0.17 | 0.07 | 0.008 |
| 55-59 | 0.17 | 0.07 | 0.008 |
| 60-64 | 0.17 | 0.07 | 0.027 |
| 65-69 | 0.29 | 0.21 | 0.027 |
| 70-74 | 0.29 | 0.21 | 0.108 |
| 75-79 | 0.29 | 0.21 | 0.108 |
| 80 and older | 0.29 | 0.21 | 0.181 |

The transmission model was implemented in C++. Data collection, data management, model runs, and results analysis and reporting were performed using R. All data and source code were bundled into an R package and a Shiny application was developed to run the model with different parameters, and to explore the various results (available upon request).

## 5 Results

### 5.1 Identification of ICU capacity limits by French Region

Our first objective was to identify when ICU capacities in each French Region would likely be overrun by the COVID-19 epidemic. Figure 4 shows the evolution of the number of required ICU beds to treat patients in critical conditions in each Region, along with the theoretical ICU capacity limits. In the worst case scenario, all Region will be overrun before April 14^*th*^, 2020. As seen on Table 2, Corse is likely to be at full ICU capacity before the end of March 2020 (March 28 in the best case, March 18 in the worst case scenario). In the scenario with *R*_0_ = 2.25, half of the French Regions will run out of ICU capacities before mid-April.

**Table 2:** Estimated date of ICU capacity overrun in the 13 metropolitan French Regions, for *R*_0_ 1.5, 2.25, and 3. Simulations start on March 10, and end on April 14, 2020.

| Region | $R_0$ 1.5 | $R_0$ 2.25 | $R_0$ 3 |
| --- | --- | --- | --- |
| Corse | 2020-03-28 | 2020-03-21 | 2020-03-18 |
| Grand-Est |  | 2020-03-28 | 2020-03-22 |
| Bourgogne-Franche-Comte |  | 2020-04-01 | 2020-03-24 |
| Bretagne |  | 2020-04-06 | 2020-03-27 |
| Hauts-de-France |  | 2020-04-07 | 2020-03-28 |
| Auvergne-Rhone-Alpes |  | 2020-04-10 | 2020-03-30 |
| Ile-de-France |  | 2020-04-14 | 2020-04-01 |
| PACA |  |  | 2020-04-05 |
| Normandie |  |  | 2020-04-08 |
| Pays de la Loire |  |  | 2020-04-08 |
| Nouvelle-Aquitaine |  |  | 2020-04-09 |
| Occitanie |  |  | 2020-04-09 |
| Centre-Val de Loire |  |  | 2020-04-12 |

**Figure 4:**
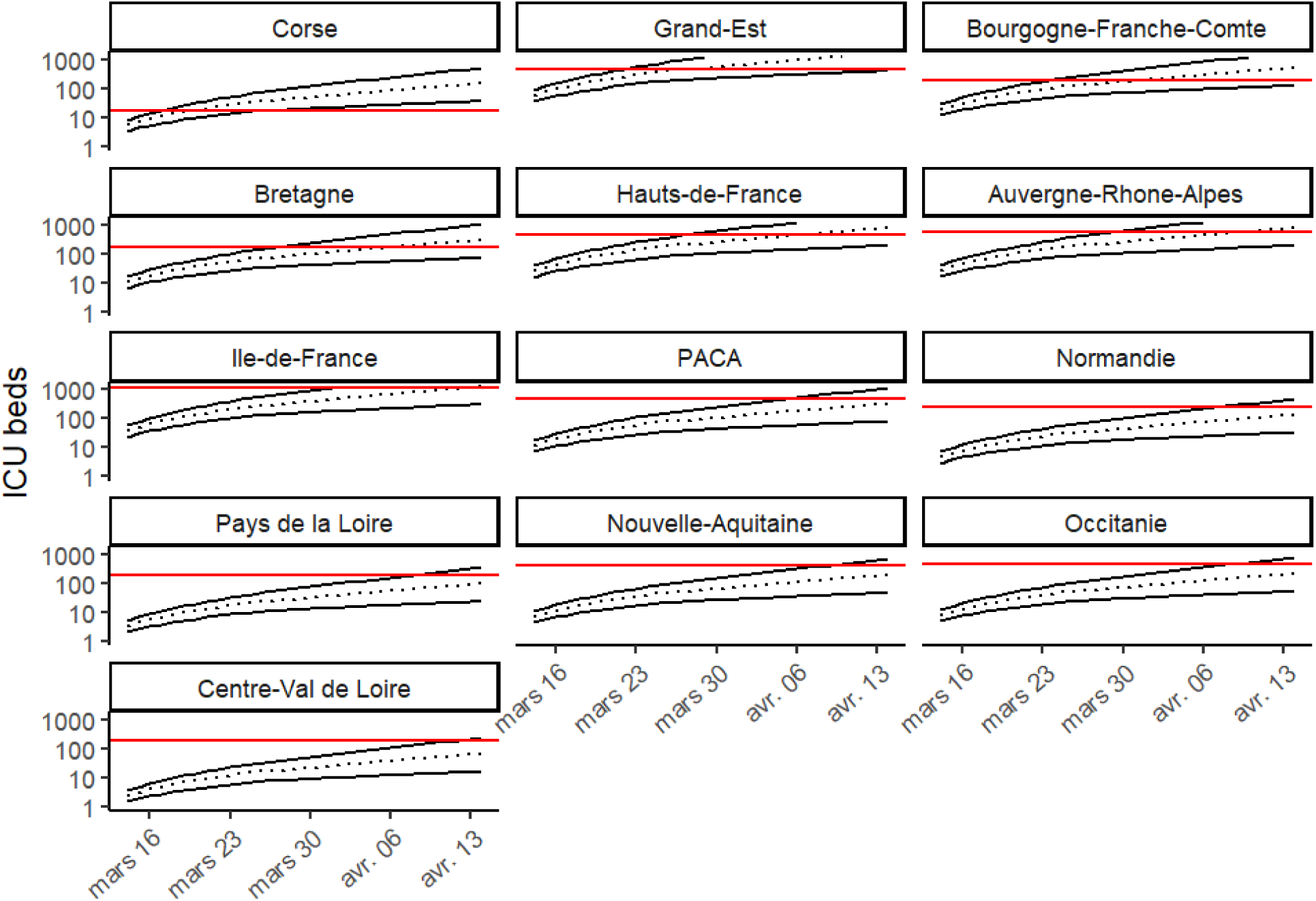
Predicted needs of ICU beds in the 13 French Regions. The red line stands for the ICU capacity limit, the dotted line stands for the scenario with *R*_0_ = 2.25, the black lines for the worst and best case scenarios (*R*_0_ = 3 and *R*_0_ = 1.5, respectively). Panels for each French Region are ordered by time of overrun (left to right and top to bottom).

### 5.2 Predicted outcomes by Region

Our analysis predict that the most affected French Region would be “Grand-Est”, with up to 42,000 infections, 10,000 severe cases and more than 2,800 deaths between March 10 and April 14 in the worst case scenario. In the same scenario, Region “Ile-de-France” and “Auvergne-Rhones-Alpes” would be the second and third most affected with up to 2,000 and 1,300 deaths, respectively. In the middle range scenario (*R*_0_ = 2.25) and best case scenario (*R*_0_ = 1.5), the morbidity and mortality burden would represent, respectively, roughly 30% and 10% of those numbers.

Our analysis shows that in France between 2,500 and 25,000 patients may require an ICU stay, and between 1,800 and 18,000 may need to be ventilated. Table 4 shows the estimated total number of ICU beds and ICU ventilated beds required per Region on April 14 under the three *R*_0_ scenarios, as well as the total ICU capacity in the Region, considering an average length of stay of 15 days. We note very large variations between the two extreme scenarios (from 305 to 4,260 ICU beds in Ile-de-France), and that the middle range scenario generally shows estimations very close or over the regional ICU capacity limit, which will restrain possible inter-regional cooperation or patient transfers.

**Table 3:** Predicted number of infected cases, severe cases, and deaths, from March 10 to April 14, 2020, by Region, for *R*_0_ values of 1.5, 2.25, and 3.

| Region | Infected |  |  | Severe cases |  |  | Deaths |  |  |
| --- | --- | --- | --- | --- | --- | --- | --- | --- | --- |
| | $R_0$ 1.5 | $R_0$ 2.25 | $R_0$ 3 | $R_0$ 1.5 | $R_0$ 2.25 | $R_0$ 3 | $R_0$ 1.5 | $R_0$ 2.25 | $R_0$ 3 |
| Auvergne-Rhone-Alpes | 2714 | 7352 | 19306 | 545 | 1624 | 4692 | 124 | 397 | 1353 |
| Bourgogne-Franche-Comte | 1851 | 5044 | 13280 | 374 | 1117 | 3233 | 86 | 275 | 934 |
| Bretagne | 1029 | 2805 | 7415 | 208 | 621 | 1806 | 48 | 153 | 522 |
| Centre-Val de Loire | 229 | 624 | 1654 | 46 | 138 | 403 | 11 | 34 | 117 |
| Corse | 536 | 1450 | 3764 | 108 | 321 | 916 | 25 | 79 | 264 |
| Grand-Est | 5984 | 16171 | 41973 | 1202 | 3571 | 10191 | 274 | 873 | 2935 |
| Hauts-de-France | 2706 | 7319 | 19105 | 542 | 1613 | 4637 | 122 | 393 | 1334 |
| Ile-de-France | 4418 | 11851 | 30754 | 879 | 2602 | 7448 | 196 | 630 | 2137 |
| Normandie | 472 | 1286 | 3403 | 95 | 285 | 828 | 22 | 70 | 239 |
| Nouvelle-Aquitaine | 682 | 1866 | 4960 | 138 | 414 | 1209 | 32 | 102 | 350 |
| Occitanie | 775 | 2112 | 5594 | 156 | 468 | 1362 | 36 | 115 | 394 |
| Pays de la Loire | 346 | 941 | 2484 | 70 | 208 | 604 | 16 | 51 | 174 |
| PACA | 1128 | 3073 | 8139 | 228 | 681 | 1982 | 53 | 168 | 574 |
| All | 22872 | 61896 | 161832 | 4590 | 13663 | 39311 | 1043 | 3339 | 11328 |

**Table 4:** Predicted number of required intensive care units beds on April 14 2020, by Region, for *R*_0_ values of 1.5, 2.25, and 3.

| Region | ICU beds |  |  | ICU beds with ventilation |  |  | Total ICU capacity |
| --- | --- | --- | --- | --- | --- | --- | --- |
| | $R_0$ 1.5 | $R_0$ 2.25 | $R_0$ 3 | $R_0$ 1.5 | $R_0$ 2.25 | $R_0$ 3 | |
| Corse | 37 | 147 | 518 | 26 | 105 | 368 | 18 |
| Grand-Est | 414 | 1645 | 5782 | 294 | 1169 | 4111 | 465 |
| Bourgogne-Franche-Comte | 128 | 515 | 1843 | 91 | 366 | 1310 | 198 |
| Bretagne | 71 | 286 | 1032 | 51 | 204 | 734 | 162 |
| Hauts-de-France | 187 | 745 | 2646 | 133 | 530 | 1882 | 438 |
| Auvergne-Rhone-Alpes | 188 | 749 | 2682 | 134 | 533 | 1907 | 559 |
| Ile-de-France | 305 | 1205 | 4260 | 217 | 857 | 3029 | 1147 |
| PACA | 78 | 314 | 1134 | 56 | 223 | 806 | 460 |
| Normandie | 33 | 131 | 474 | 23 | 93 | 337 | 240 |
| Pays de la Loire | 24 | 96 | 346 | 17 | 68 | 246 | 181 |
| Nouvelle-Aquitaine | 47 | 191 | 692 | 34 | 136 | 492 | 412 |
| Occitanie | 54 | 216 | 780 | 38 | 153 | 554 | 474 |
| Centre-Val de Loire | 16 | 64 | 231 | 11 | 45 | 164 | 180 |

## 6 Discussion

In the context of few available modeling data on COVID-19 epidemic in France, the purpose of our study was to provide a one-month forecast of the spreading dynamic of the virus across all metropolitan French Regions, and to assess hospitals burden, especially regarding ICU beds and ventilation needs.

We modeled the propagation of COVID-19 from March 10 to April 14, across all metropolitan French Regions. At the national level, the total number of infected cases is expected to range from 22,872 in the best case (*R*_0_ = 1.5) to 161,832 in the worst considered case (*R*_0_ = 3). Regarding the total number of deaths, it is expected to vary from 1,021 to 11,032, respectively. At the regional level, all ICU capacities may be overrun in the worst scenario, Centre-Val de Loire being the last region to reach its limits on April 12. Only seven Regions may lack ICU beds in the mid scenario (*R*_0_ = 2.25) and only one in the best case. In the three scenarios, Corse may be the first Region to see its ICU capacities overrun with a delay of one week between each scenario, which may be due to its low ICU capacity (18 beds). The two other Regions, whose capacity will be overrun shortly after, are Grand-Est and Bourgogne-Franche-Comté, despite their high number of ICU beds (465 and 198, respectively).

COVID-19 mortality risk is not only dependent on the intrinsic virus’ characteristics, but depends also on health care capacities, especially ICU. Italy is already seeing its ICU capacities exceeded, as well as several hospitals in France. However, few studies have tried to forecast ICU needs. Grasselli and colleagues used linear and exponential models to estimate ICU demand in Lombardy, Italy, from March 7 to March 20^28^. They concluded that the health care system could not sustain an uncontrolled outbreak, and that strong containment measures were needed. Remuzzi and Remuzzi fitted exponential models and predicted that the Italian health care system could be at maximum capacity by March 14^29^. Here, we found that ICU capacities would be rapidly exceeded in several Regions in France, even in optimistic scenarios. Incapacity to provide adequate treatment to critically ill patients could further increase the already substantial death toll of the epidemic.

Besides limits inherent to each transmission model, this work has several specific limitations. The model was run for each catchment area independently, as we did not model population movements between catchment areas. Although it is theoretically feasible, it seems unnecessary in this context. Infected cases are already present in most locations, meaning transmission is likely to be mainly driven locally, not by inter-areas transfers. Moreover, control measures are already implemented to limit population movements. We are only presenting forecasts at one month, as long term predictions may be unreliable due to the few data available to calibrate the model concerning the epidemic in France. The critical factor that remains unknown to this date is the potential impact of seasonality on the transmission dynamic of COVID-19. Danon and colleagues modeled seasonal transmission by introducing a time-varying transmission rate.^21^ They estimated that a 50% reduction in transmission during summer months would result in a smaller epidemic before the summer, followed by a resurgence in cases in the following winter. However, whether SARS-CoV-2 transmission will be affected by seasonal variations remains unclear. Although many infectious diseases have seasonal patterns, like influenza or other coronaviruses, newly introduced viruses can behave differently. Several experts suggest that the impact of seasonality on COVID-19 transmission might be very modest,^30,31^ therefore we did not included such a seasonality in our model at the time. Also, we decided to explore only *R*_0_ values between 1.5 and 3, as it seems to be the most reliable estimates at the moment, according to Abbott and colleagues^20^. Finally, we did not make a distinction between symptomatic and asymptomatic cases, whereas some modelers decided to create two compartments (infectious asymptomatic and infectious symptomatic). We believed it was unnecessary in this context as we are not focusing on detection or control measures, and it should not impact the interpretation of our results.

Regarding the prediction of ICU needs, the ICU capacities used in this study are theoretical and based on annual data,^24^ and may not reflect exactly current ICU capacities. However, the potential discrepancies are likely to be modest and should not impact the interpretation of the results. More importantly, we assumed that ICU capacities could entirely be dedicated to COVID-19 critical patients. In France, ICU wards tend to have more than 80% occupancy rate (personal communication with clinicians), hence our analysis already assumes an important patient care reorganization, almost doubling current capacities. Also, we assumed a fixed length of stay of 15 days in ICU as observed in foreign countries. This value could vary across patients and locations, and could slightly bias the estimation of ICU needs in our study. To overcome this bias, we plan to use hospitalization data provided by regional health agencies (ARS), which may help to have better predictions of beds use.

We restricted our model to metropolitan France, as overseas Regions are at a lower epidemic stage at the moment, and the transmission dynamics are likely to differ from metropolitan France. Finally, it must be noted that spatial division of the French territory used in this model does not exactly reflect administrative boundaries of the French Regions, as it was based on the merge of each Voronoi polygons representing the catchment areas of referral centers. However, division by catchment areas might help to better estimate the pressure on the referral centers, as people are more likely to be referred to hospitals based on geographical distance rather than administrative boundaries.

To mitigate the epidemic, the French government announced a series of nation-wide measures, of increasing importance. On February 29, gatherings were restricted to 5,000 persons in confined settings. This threshold has since been revised downwards, with 5,000 persons on March 8, and 100 persons on March 13. On March 12, schools and universities were asked to close starting from March 16, and voluntary household quarantine and social distancing of those over 70 years of age were recommended. Finally, on March 14, the stage 3 of the epidemic was declared. Consequently, all non-essential public places (i.e. non-food shops, restaurants, pubs,…) are closed, and people are enjoined to stay home, limit social interactions, and travels as much as possible. These mitigation measures were not directly implemented in the present study, but their impact was assessed indirectly by modifying the value of the reproduction number.

## 7 Conclusion

While preliminary, our analysis shows that, even in the best case scenario, the French healthcare system will very soon be overwhelmed. While drastic social distancing measures may temper our results, a massive reorganization leading to an expansion of French ICU capacities seems to be necessary to manage the coming wave of critically affected COVID-19 patients.

## Data Availability

All data and source code are available upon request.

